# Investigating the contributions of circadian pathway and insomnia risk genes to autism and sleep disturbances

**DOI:** 10.1101/2021.06.24.21259489

**Authors:** Rackeb Tesfaye, Guillaume Huguet, Zoe Smiliovich, Mor Absa Loum, Elise Douard, Martineau Jean-Louis, Jean Luc Martinot, Rob Whelan, Sylvane Desrivieres, Andreas Heinz, Gunter Schumann, Caroline Hayward, Mayada Elsabbagh, Sebastien Jacquemont

## Abstract

Sleep disturbance is prevalent in youth with Autism Spectrum Disorder (ASD). Researchers have posited that circadian dysfunction may contribute to sleep problems or exacerbate ASD symptomatology. However, there is limited genetic evidence of this. It is also unclear how insomnia risk genes identified through GWAS in a general population are related to ASD risk and common sleep problems like insomnia in ASD. We investigated the contribution of copy number variants (CNVs) encompassing circadian pathway genes and insomnia risk genes to ASD risk as well as parent reported sleep disturbances in children diagnosed with ASD. We studied 5860 ASD probands and 2092 unaffected siblings from the Simons Simplex Collection and MSSNG database, as well as 7463 individuals from two unselected populations (IMAGEN and Generation Scotland). We identified 320 and 626 rare CNVs encompassing circadian genes and insomnia risk genes respectively. Deletions and duplications with circadian genes were overrepresented in ASD probands compared to siblings and unselected controls. For insomnia-risk genes, deletions (but not duplications) were also associated with ASD. Results remained significant after adjusting for cognitive ability. CNVs containing circadian pathway and insomnia risk genes showed a stronger association with ASD, compared to CNVs containing other genes. Duplications containing circadian genes were associated with shorter sleep duration (22 minutes). Only insomnia risk genes intolerant to haploinsufficiency increased insomnia traits when duplicated. Overall, CNVs encompassing circadian and insomnia risk genes increase ASD risk despite small impacts on sleep disturbances.

## Introduction

Sleep problems have been reported in 40 to 80% of youth with Autism Spectrum Disorder (ASD)[1–4] These problems emerge early in infancy and continue throughout the lifespan among those diagnosed with ASD[5]. The most commonly reported sleep disturbances in ASD include poor sleep duration and insomnia, defined by night awakenings and/or delayed sleep onset that contribute to negative daytime functioning consequences. Sleep is crucial for successful mental and physical well-being, and such disturbances may contribute to exacerbating symptomatology and comorbidities in ASD [6]. However, the biological processes underlying elevated sleep problems in ASD remains understudied. Such mechanisms may be informed by genetic factors, which are major contributors to ASD liability as well as sleep disturbance. Heritability of ASD is between 64 – 90%[7], while the heritability of self-reported sleep duration and insomnia in adults ranges between 0.25%-0.44% and 0.22%-0.59% respectively[8, 9].

The daily timing of sleep, in addition to other physiological functions (i.e., behavioral, hormonal, and attentional processes), is regulated by the circadian system[10]. It’s composed of an oscillatory rhythm that fluctuates to an approximate 24-hour period. Circadian rhythms are generated in the suprachiasmatic nucleus are regulated by “clock” genes and influenced by environmental cues such as light, temperature and social activities[10, 11]

Earlier genetic studies of sleep traits tested the hypothesis that mutations in clock genes would affect circadian sleep phenotypes[9, 12]. For instance, common variants in *CRY1* were associated with delayed sleep phase disorder[13], while variants in *PER2[14, 15]* and *CKIδ*[16] led to advanced sleep phase disorders.

Researchers have put forth a circadian theory of ASD risk, suggesting that circadian dysfunction may underlie elevated sleep problems, which increases the susceptibility of an ASD diagnoses, along with other difficulties related to circadian misalignment (e.g., social cues, attentional processes, etc.)[17–19]. The link between ASD and circadian pathways has been investigated in a few studies, however, the potential relationship remains unclear. Studies, mainly focusing on clock genes, were not able to associate common variants in circadian genes with ASD or sleep disturbances in this population[20–23]. As an example, a 2019 study showed SNPs within 25 clock and melatonin genes were not associated with broad night and daytime sleep issues in 2065 ASD youth in the Simon Simplex Collection[22]. The relationship between the circadian system and ASD has also been investigated through melatonin regulation. Atypical levels of melatonin (neurohormone that helps reset the biological clock[24]) have been found in ASD and studies suggest that common genetic variants altering melatonin synthesis are associated with sleep phenotypes in youth with ASD[17, 25, 26].

However, most of the genetic contribution and biological processes implicated in sleep disturbances appear to lie outside the circadian pathway. This is demonstrated by a recent genome wide association study on insomnia that identified only a handful of circadian genes among the 956-insomnia risk-genes identified[27]. Further, genetic overlaps were found between insomnia and neuroticism, depressive symptomology, and schizophrenia, in addition to other traits, but not ASD[27].

Presently, it is unclear whether larger circadian pathway and insomnia risk genes contribute to ASD risk and/or to sleep problems in individuals with ASD. In particular, there has been no focus on rare variants implicating circadian and insomnia-risk genes. The importance of rare copy number variants (CNVs; defined as genomic deletions or duplications >1kb) to ASD is well demonstrated and previous studies have replicated the association between 16 specific recurrent CNVs and ASD [28]. Our team has also shown that rare non-recurrent CNVs distributed across the genome encompassing coding genes intolerant to haploinsufficiency are associated with increased liability to ASD[29]. Whether circadian and insomnia-risk genes encompassed in CNVs contribute to ASD risk is unknown. In addition, the effects of these CNVs on sleep problems have yet to be investigated in either a general population or ASD population.

We hypothesize that genomic variants disrupting pathways involved in circadian rhythms and insomnia are linked to ASD risk. Our aim was to understand the relationship between autism risk, sleep disturbances, circadian pathways and insomnia-risk genes.

We investigated circadian and insomnia risk genes disrupted by CNVs in two ASD cohorts (Simons Simplex Collection and MSSNG), and in unaffected siblings (Simons Simplex Collection) and individuals from unselected populations (Generation Scotland and IMAGEN). We also characterized the effect of these CNVs on parent reported sleep duration and insomnia traits.

## Materials and Methods

### Cohorts

#### Autism Datasets

The Simons Simplex Collection (SSC) includes 2,569 simplex families with one ASD proband per family and 2,851 unaffected siblings. The MSSNG dataset, was used as an independent replication cohort, and includes 3,426 probands with ASD from multiplex families. [30–32].

#### General Population

The general population was pooled from two previously described cohorts. The longitudinal IMAGEN dataset containing 2,093 adolescents[33]. The second cohort included 16,916 adults from Generation Scotland: the Scottish Family Health Study (GS) [34]. Only one individual per family was included.

### CNV calling, filtering and annotation

In all cohorts except MSSNG, SNP array data was available and CNVs were called using Penn CNV and QuantiSNP based on published pipelines [35]. For MSSNG, CNV’s were called from whole genome sequencing using Trost et al., published pipeline[36].

CNVs were annotated using ANNOVAR and the UCSC Genome Browser to map segmental duplications, centromeric and HLA regions. We annotated genes encompassed in the CNVs using Gencode V19 annotation (the reference release for hg19 Human genome release) with ENSEMBL(https://grch37.ensembl.org/index.html).

Rare CNVs in each cohort were defined by: a*) frequency* < 1/1000 in Data Genome Variants (DGV, hg19, http://www.dgv.tcag.ca) with an overlap of 70%; and < 1/1000 of the population frequency (individuals within the same cohort served as the population control); and b) the CNV is contained in regions present at > 1% in DGV< 50%.

We selected CNVs ≥ 50Kb; with less that 50% overlap with segmental duplications, centromeric or HLA regions. We only analyzed autosomal CNVs since the effect of gene dosage is not comparable between sex-linked and autosomal CNVs.

For harmonization we selected CNVs encompassing at least ten probes for all array technologies used across all cohorts.

### Sleep Risk Gene Lists

*Circadian pathway* genes (n=334) were identified in three genetic databases: KEGG Pathway (hsa04710/hsa04713) [37], GeneOntology(GO:0007623) [38], and REACTOME[39] (R-HSA-1368108/R-HSA-400253). Gene sets were extracted in May 2019. Core clock genes within this list were identified using existing literature[11].

*Insomnia risk* genes (n=953) were extracted from the largest GWAS on insomnia[27]. All identified genes were filtered out if they were non-protein coding or pseudogenes based on published datasets from The Human Genome Organization[40].

High confidence ASD-risk genes were collected from the SFARI Gene database (category 1 and 2) [41]. Overlaps between extracted lists of ASD risk, circadian pathway and insomnia risk genes are found in Figure1.

### Scores of intolerance to haploinsufficiency and brain expression

All CNVs were scored based on the LOEUF values of genes encompassed in CNVs were scored *using “*loss-of-function observed/expected upper bound fraction” (LOEUF) as previously published[42]. For each CNV, we computed the sum 1/LOEUF of all genes encompassed in that CNV, hence a high CNV score indicates a strong intolerance toward inactivation. The differential stability (DS) score[43] is a correlation-based metric that assess reproducibility of spatial patterns of gene expression in the brain. The score was transformed and ranges between 0 and 1, where a higher score indicates stable gene expression in specific brain regions. We computed the sum of DS scores of all genes encompassed in each CNV.

### Clinical and behavioral data

*The SSC Sleep Interview (SSCI)* is an 11-item parent report questionnaire assessing nighttime and daytime problems and sleep duration problems[44] see Supplemental Materials. Sleep data was only available for the SSC ASD probands. For the purposes of our study, we analyzed two sleep traits commonly reported as disturbed in ASD:

1. Sleep duration: Average duration of sleep per week in minutes
2. Insomnia corresponding to two items: ‘*difficulty falling asleep’* and *‘frequent or prolonged awakenings at night*. These items parallel self-reports used to measure insomnia in Jansen et al.,[27]. The number of insomnia traits was scored from 0-2.

Other binary sleep traits, including daytime sleepiness and troubles waking up in the morning were extracted for descriptive purposes.

#### Cognitive ability

Non-verbal IQ (NVIQ) data were available for the ASD cohorts and the IMAGEN dataset. A *g* factor was available for GS. Cognitive assessment methods for each cohort are detailed in *Supplementary Materials*. No cognitive information was collected for unaffected siblings.

*The Autism Diagnostic Observation Scale calibrated severity score (ADOS CSS)[45]* is a 10-point scale based on raw ADOS scores. It captures overall ASD symptom severity independent of age and language level. Higher scores indicate greater symptom severity.

### Data Analysis

All analyses were performed with R 3.6.3 (Supplementary Materials).

### Association between ASD and circadian and insomnia risk genes

Bayesian logistic regression analysis was used to estimate this association as follows:

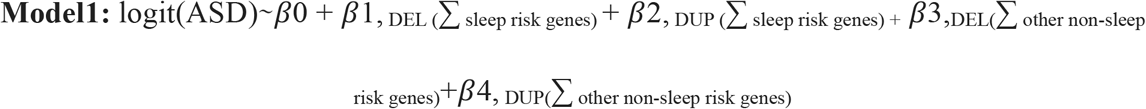

*DEL= Deletion; DUP=Duplication*

where 0 is the intercept and 1-3 are the regression coefficients. Other non-sleep genes refer to protein coding genes in the genome that are non-circadian pathway and non-insomnia risk genes, encompassed in a CNV.

#### Secondary analyses

Follow up logistic regression analyses investigating the effects of gene intolerance (1/LOEUF) and brain expression (DS_Score_)were also conducted.

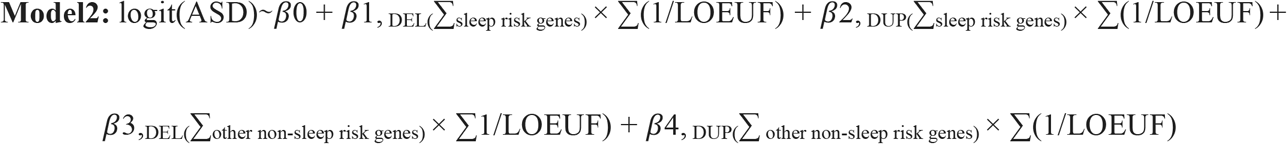

1/LOUEF scores per gene was summed by type of CNV (DEL/DUP) and gene category (sleep/non-sleep risk genes). This same model was used with DS scores.

For all models above, two additional identical models were performed separating CNV’s encompassing circadian and insomnia genes. Models comparing ASD probands and unaffected siblings are adjusted for familial relationship with a random effect. Follow up models above were controlled for cognitive ability.

#### Bootstrapping

To test how robustly CNVs encompassing selected sleep risk genes better-predicted autism risk, we compared 95% confidence intervals obtained using a bootstrap procedure (1,000 iterations) between sleep and non-sleep gene duplications and deletions.

### Associations between sleep genes and SSC sleep phenotype

A linear regression was applied to test the association between sleep duration in minutes and sleep risk genes.

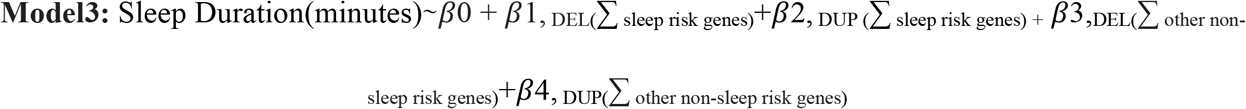

An ordinal logistic regression was used to analyze the association between sleep risk genes and the number of insomnia traits (three levels: 0, 1, 2).

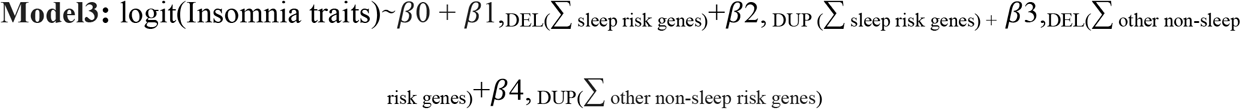

Relevant co-variates were added to all sleep phenotype models above (Supplementary Materials). Additional analyses to determine the association between sleep phenotypes and the sum of 1/LOEUF and DS_score_ per CNVs were conducted. All models were followed up by separating CNV’s encompassing circadian and insomnia genes.

### Multiple comparisons

Bonferroni corrections were applied for each main model to account for our three genetic constraint scores tested (i.e., binary,1/LOEUF, DS_score_). Significance was corrected to p=0.0167, subsequent sensitivity analyses were not corrected for.

## Results

### Associations between sleep (circadian and insomnia) risk genes and autism risk

Demographic data is found in Table 1. Overall, we identified 249 duplications and 72 deletions encompassing circadian pathway genes, as well as 376 duplications and 250 deletions encompassing insomnia risk genes (Table 1). Of note, there was minimal overlap between our list of sleep risk genes and the SFARI list of high confidence ASD risk genes (Figure 1, Supplementary Table 1-6).

**Table. 1.**
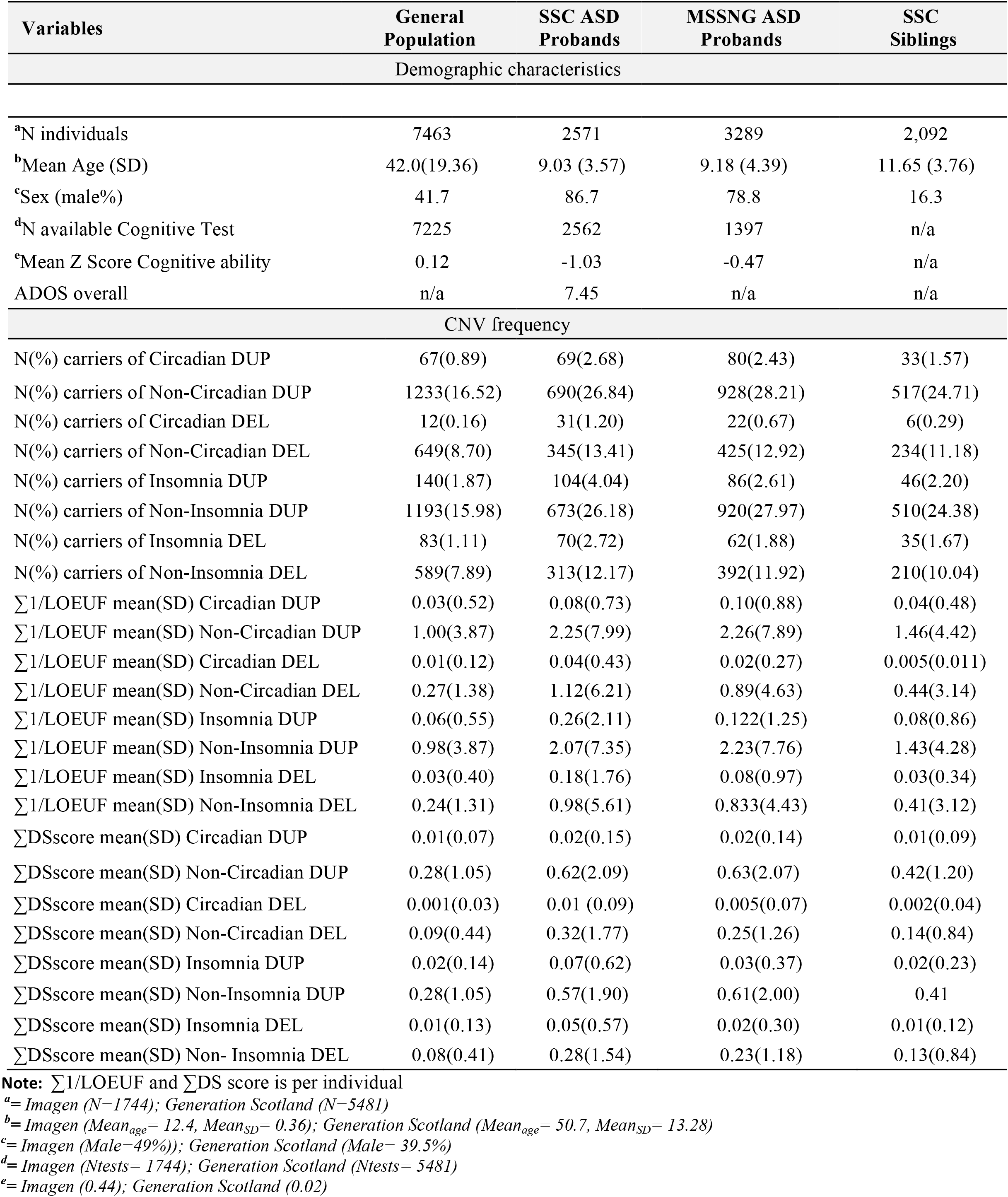
Cohort descriptives

**Figure 1.**
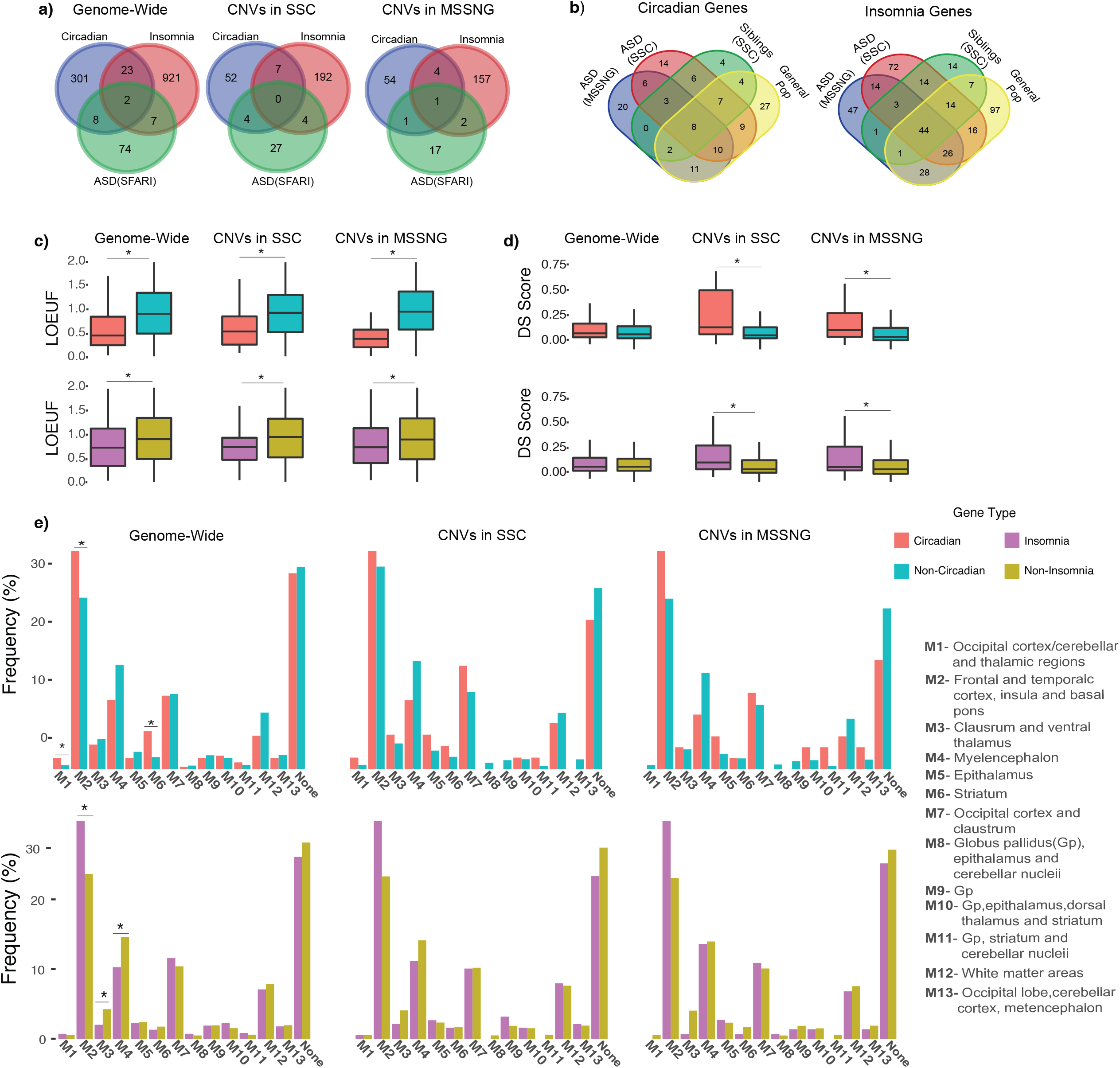
Sleep risk gene characteristics. a) Overlaps between insomnia risk, circadian pathway and ASD risk genes. b) Overlap between insomnia risk and circadian pathway genes encompassed in CNV’s in each cohort. c) LOEUF score comparisons between insomnia risk and circadian pathway genes. Mann Whitney-U tests applied. d) DS score comparisons between insomnia risk and circadian pathway genes. Mann Whitney-U tests applied. e) Brain module (Hawrylycz et al., 2021) comparisons between insomnia risk and circadian pathway genes. Modules provide the locations of gene brain expression (see legend). Binomial linear regressions were used to compare modules and FDR corrections were applied. * = signifigant

#### Deleting or duplicating sleep risk genes is associated with autism risk

##### SSC ASD probands vs general population

Rare duplications (OR = 1.75, *p* = 3.34^-03^) and deletions (OR = 1.79, *p* = 9.00^-03^) encompassing any sleep risk gene were significantly overrepresented in SSC probands. This remained after subdividing into circadian (OR for duplication = 2.01, *p =*6.67^-05^ and deletions = 4.83, *p* = 4 .01^-06^) as well as insomnia risk-genes (OR for duplications = 1.60, *p* = 5.91^-04^ and deletions = 2.16, *p* = 3.63^-06^).

##### SSC ASD probands vs Unaffected Siblings

The same analysis using unaffected siblings as a control group provided similar results with an overall weaker signal. No differences of CNVs encompassing circadian and insomnia risk genes were found between unaffected siblings and the unselected population.

##### MSSNG ASD probands vs general population

Enrichments similar to SSC were observed in the MSSNG cohort: (OR for deletions and duplications of circadian genes =3.24, *p* = 1.02^-03^ and 1.77, *p* = 7.84^-04^ respectively, as well as OR for deletion of insomnia genes =1.53, *p* = 1.26^-02^) except for duplications of insomnia genes. Pooling the MSSNG and SSC datasets yielded similar results.

For results above see Supplementary Table 7 and Figure 2.

**Figure 2.**
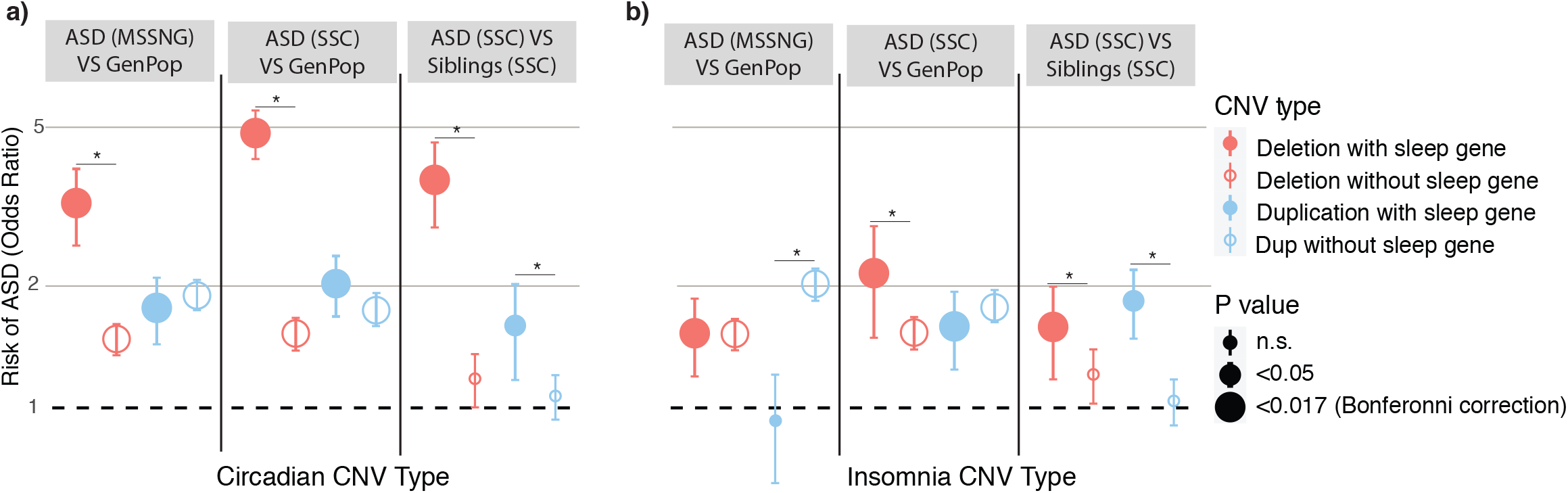
Associations between ASD and sleep risk genes. a) Associations between ASD risk and CNVs encompassing circadian pathway genes. b) Associations between ASD risk and CNVs encompassing insomnia risk genes. * indicates bootsrap analyses show no overlapping 95% CI

Sensitivity bootstrap analyses confirmed results above. Additional analysis showed that deletions containing circadian and, to a lesser extent, insomnia risk genes were better predictors of ASD risk compared to deletions encompassing other genes (Supplementary Table 7). Effects of circadian genes but not insomnia-risk genes remained significant after controlling for IQ in a pooled SSC / MSSNG dataset (Supplementary Table 8). Excluding all recurrent neuropsychiatric CNVs[35] demonstrated similar enrichments of circadian and insomnia-risk genes in the pooled dataset.

#### Insomnia-risk genes intolerant to haploinsufficiency increase ASD risk

Based on previous studies[29], it was expected that autism risk uncovered above would be driven by genes intolerant to haploinsufficiency. Circadian and insomnia genes showed a distribution of LOEUF values significantly skewed towards intolerance (Figure 1). For insomnia genes, ASD risk was associated with increasing intolerance to haploinsufficiency measured by LOEUF in the pooled dataset for duplicated (OR= 1.08, *p* = 1.15^-03^) and deleted (β= 1.11, *p* = 8.47^-03^) insomnia genes compared to the general population (Supplemental Table 9). However, measures of intolerance did not influence autism risk conferred by circadian genes.

#### Brain expression of sleep genes is associated with autism risk

We expected that circadian and insomnia risk genes with stronger patterned brain expression (measured by DS score [43]) would be related to autism risk. The full list of circadian genes show a slightly higher DS score compared to non-circadian genes (Figure 1). This was not the case with the insomnia gene list. However circadian and insomnia risk genes encompassed in CNVs identified in both ASD cohorts had a slightly higher DS score compared to non-sleep genes (Figure 1).

In both cohorts, ASD risk was related to higher DS scores for circadian genes encompassed in duplications (OR= 1.80, *p* = 4.76^-03^). This was not found for circadian deletions or insomnia risk genes. Supplementary Table 10.

### Associations between CNVs and sleep phenotypes

#### Descriptives

Sleep duration and insomnia traits were available for 2532 and 2473 SSC probands respectively (Table 1). Mean sleep duration adjusted for age was 660 minutes (Figure3). Age and NVIQ, were weakly associated with sleep duration, while sex and ADOS overall severity were not (Supplementary Table 11). 39% of individuals in SSC had at least one insomnia trait. NVIQ and sex (i.e., females had more insomnia) were the only covariates associated with insomnia traits -albeit weakly (Supplementary Table 12). Probands with two insomnia traits were twice as likely to experience daytime sleepiness (37%) and difficulties waking up in the morning (29%) compared to the rest of the cohort (14 and 17% respectively; Supplementary Table 13). These associations demonstrate the negative impact insomnia traits have on daily functioning.

**Figure 3.**
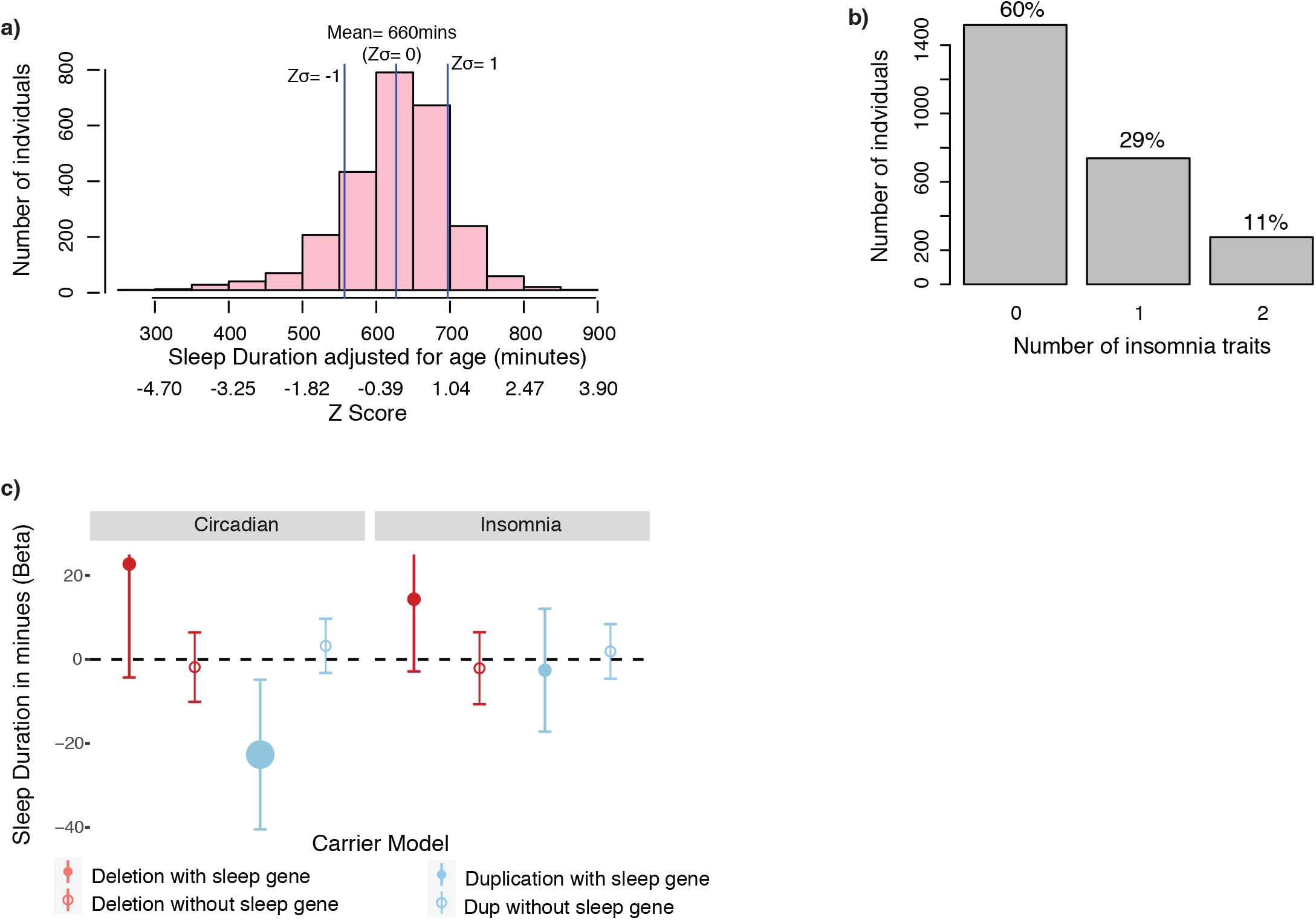
Sleep trait descriptions and gene associations in the SSC ASD cohort. a) Sleep duration distribution adjusted for age. b) Frequency of insomnia traits. c) A linear regression reveals ASD probands with a duplication eompassing a circadian pathway gene (left in blue fill) have 22 minutes of decreased sleep duration.

#### Circadian duplications decrease sleep duration

Only duplicated circadian genes decreased sleep duration. ASD youth with a rare circadian gene duplication showed a decrease of 22 minutes (0.34 z-score) per weeknight compared to ASD youth without such duplications (p=0.013) (Supplementary Table 14). To further investigate the effects of circadian gene duplications, we removed 86 individuals with recurrent neuropsychiatric CNVs. Findings were unchanged, with an average loss of 24 minutes of sleep per weeknight (*p* = 0.03; Supplementary Table 15). However, we did not observe any differences in parental report of daytime sleepiness or difficulties waking up in the morning in the group with circadian duplications compared to non-carriers.

In contrast, deletions (but not duplication) encompassing non-circadian and non-insomnia genes scored by LOEUF or DS, led to an increase in sleep duration (Supplementary Table 14).

#### Insomnia genes have mild effects on insomnia traits

The presence of a CNV encompassing a circadian or insomnia risk gene was not associated with an insomnia trait.

However, insomnia genes measured by LOEUF scores (measuring intolerance to haploinsufficiency) were associated with increasing the likelihood of having insomnia traits when duplicated (albeit with a small effect, OR= 1.05 p = 8.33^-03^) (Supplementary Table 16). When individuals with psychiatric CNVs were removed, these findings became marginally significant (p=0.05; Supplementary Table 17).

Conversely, duplications encompassing non-insomnia genes and non-circadian genes were associated with fewer insomnia traits. The DS score of sleep risk and non-sleep risk genes was not significantly associated with insomnia traits (Supplementary Table 16).

## Discussion

We investigated the effects of CNVs encompassing circadian and insomnia risk genes on ASD risk and sleep traits. We show that insomnia risk and circadian pathway genes increase ASD liability even after adjusting for NVIQ, although these finding were less robust for insomnia duplications. Circadian pathway deletions were better indicators of ASD risk compared to deletions that did not contain circadian genes. Our results also suggest that circadian genes may alter sleep duration, without influencing insomnia traits. On the other hand, insomnia risk genes were associated with insomnia traits without altering sleep duration. In contrast, duplications of non-sleep risk genes were associated with less insomnia traits, and deletions of non-circadian genes (measured by intolerance scores) increased sleep duration.

### Circadian pathway genes increase ASD risk and decrease sleep duration without affecting insomnia traits

It is well established that dysregulation of circadian rhythms (e.g. shift workers, sleep phase disorders) is linked with a host of psychiatric disorders, medical conditions and cognitive and behavioral impairments[46]. However, candidate SNP studies are unable to provide robust evidence of associations between circadian clock genes and psychiatric disorders[47, 48].

Our findings provide evidence for the contribution of circadian pathway dysfunction to ASD, but the mechanisms by which these genes increase ASD risk are unknown. Previous theories have put forth sleep disturbance as a causal factor for ASD risk and other psychopathologies. These theories draw on evidence from mouse models indicating that sleep disturbances early in life can lead to alterations of plasticity, brain maturation, and organization[17]. However, in this study, the limited effects of circadian genes on sleep duration and insomnia are unlikely to disrupt behavior and cognition enough to increase ASD risk.        Instead, we propose that circadian pathway genes increase ASD risk and independently modulate sleep duration. Alternatively, it is also possible that these circadian gene mutations are responsible for significantly disturbing sleep at the electrophysiological level, which may not be captured by parent reports. Such discordance between atypical electrophysiological sleep and normal sleep reported by parents in youth with ASD has previously been reported[49]. Further investigations including objective physiological sleep measures are warranted.

### Insomnia risk genes increase ASD and insomnia risk

Contrary to the largest insomnia GWAS[27] showing no genetic correlation between insomnia and ASD, we found that the gene list provided by this study increased ASD liability when deleted or duplicated (although the latter was not replicated). Although insomnia risk genes within CNVs identified in ASD cohorts showed higher patterned expression in the brain and slightly increased intolerance to haploinsufficiency compared to other coding genes in the genome, neither characteristic was clearly responsible for driving the association between insomnia risk genes and ASD. The genetic correlations between insomnia and multiple psychiatric symptoms and conditions suggest that underlying genes target mechanisms responsible for psychopathology, whereby poor sleep is one among many symptoms. As an example, the 16p11.2 deletion, which encompasses insomnia risk genes, is known to increase ASD risk and studies in mice and humans have observed circadian rhythm and sleep disturbances in those carrying this deletion [50, 51].

### Parent reported sleep disturbances are not major causal factors of ASD

Proponents of the circadian dysfunction theory suggest disturbances of circadian sleep rhythmicity may increase the vulnerability of developing ASD symptomology[17, 23]; our study did not support this. Rather, we showed that sleep problems of youth with ASD in the SSC cohort were comparable to what has previously been reported in typically developing cohorts[52, 53] Only 4% of SSC youth slept less than the National Sleep Foundation’s recommended guidelines for their age[53]. A previous SSC study reported a higher rate (∼25%) of youth that did not meet recommended sleep duration, but classified those with “may be acceptable” duration as poor sleepers [44].

Although insomnia traits in SSC were linked to greater daytime sleep consequences, reports of two insomnia traits in the SSC cohort (10%) were drastically lower than reports in typically developing youth populations[54]. These comparisons should be interpreted with caution given that insomnia traits in SSC lack information about their severity and frequency. Moreover, ASD severity, and specifiers such as NVIQ, had almost no association with sleep duration or insomnia traits. Hence, previous suggestions that sleep problems in ASD may occur from an ineffectiveness to process *environmental cues* that entrain circadian rhythms due to social and communication difficulties, were not proven in the SSC.

## Limitations

Investigating rare variants affecting gene lists representing less than 10% of the coding genome requires powerful datasets. In particular there are less than 25 known core clock genes. Slightly different results between ASD cohorts may be due to power issues and noise introduced by different technologies (microarray vs whole genome sequencing), which may contribute to discrepancies in CNV identification.

Given the limited sleep phenotypes, and in particular the absence of phenotypes in unaffected siblings and controls, we were unable to establish the normative association between sleep phenotypes and our genes of interest. Objective sleep measures are needed to validate findings between rare gene variants and sleep problems in ASD. We were not able to adjust for medication impacting sleep, which is commonly administered to ASD youth. Specifically, the use of melatonin known to ameliorate sleep problems in ASD [26], was not documented in SSC.

## Conclusion

Our results implicate rare circadian and insomnia risk genes variants with both ASD risk and sleep traits suggesting pleiotropic effects for these genes[12]. We are currently unable to compare our findings to studies of a similar or larger scale, hence further investigations in health and disease are required to delineate the phenotypic effects of circadian pathways and insomnia risk genes. We were not able to demonstrate that sleep disturbances were risk factors for ASD. Future studies investigating the combined effect of genomic variants and environmental factors on sleep measures, behavioral traits and brain architecture are needed for a holistic understanding of the interplay between genes, sleep and ASD.

## Supporting information

Suppl_Information_Tesfaye2021

SupplTables_Tesfaye2021

## Data Availability

Researchers can request data used in the current study by following established data access protocols for each cohort listed below:
Simon Simplex Cohort https://www.sfari.org/resource/sfari-base/
MSSNG https://science.grants.autismspeaks.org/genetics/login.php
IMAGEN https://imagen-europe.com/resources/imagen-project-proposal/
Generation Scotland
https://www.ed.ac.uk/generation-scotland/for-researchers/access

## Acknowledgments

We’d like to thank all individuals and families who participated.

