## Supplementary material for "Investigating the contributions of circadian pathway and insomnia risk genes to autism and sleep disturbances": Suppl_Information_Tesfaye2021

**SUPPLEMENTARY MATERIALS ONLINE CONTENT**

**METHODS**

CNV Technology

- SSC: *Microarray (*1Mv1, 1Mv3, and Omni 2.5)
- MSSNG: *Whole Genome Sequencing (*HiSeq, HiSeq 2,500, and HiSeqX)
- IMAGEN: *Microarray (*Illumina Quad 610 / 660 chips)
- Generation Scotland: *Microarray (*Illumina GSA)

CNV Detection

CNV detection algorithms from the software PennCNV[1] and QuantiSNP[2] , were used with the

following parameters: a) 3 consecutive probes for CNV detection; b) CNVs > 1 Kb; and c)

confidence scores ≥ 15. They were then combined with CNVision[3] . After this this merging step,

the CNV Inheritance Analysis algorithm (developed by the Thomas Bourgeron’ lab), was applied

to concatenate adjacent CNVs into one, according to the following criteria:  a) gap

≤150 Kb between the CNVs; b) size of the CNVs ≥ 1000 bp; and c) number of probes ≥ 3. Visual

validations were performed with SnipPeep (<http://snippeep.sourceforge.net/)>. Finally, random

forest validation was used to filter CNVs with an algorithm trained on ~ 24,000 CNV

visualizations.

*De novo* CNVs in the SSC were identified in probands, unaffected siblings, and unselected population from SYS using two previously published datasets[3, 4], combined with our own algorithm developed in R [5]. A CNV was considered as *de novo* only if it was defined as such by all three approaches.

*Psychiatric CNVs.* Recurrent psychiatric CNVs removed for sensitivity analyses were accessed from Huguet et al., 2018[6].

**Clinical and behavioral data**

Cognition.

*SSC Autism Cohort. N*on-verbal intelligence quotient (NVIQ) scores were obtained from the Differential Ability Scales, 2nd Edition (DAS-II)[7] for early years (N=1,030) and school age children (N=1,212), the Wechsler Intelligence Scale for Children, 4th Edition (WISC-IV) (27) (N=45), the Wechsler Abbreviated Scale of Intelligence – First Edition (WASI-I)[8] (N=62) or the Mullen Scales of Early Learning (MSEL)[9] (N=213). Norm-referenced standard scores (deviation NVIQ) were available for most of the participants. However, for individuals from SSC who were not able to obtain a deviation NVIQ due to their age and/or developmental level, ratio IQ were derived by dividing mental age by chronological age and multiplying by 100. See Bishop et al., 2011 for more details concerning convergence between ratio and deviation NVIQ[10].

*MSSNG Autism Cohort.* NVIQ scores were obtained from the Leiter international performance scale – Original and revised[11,12](N=372), the raven progressive matrices[13](N=214), the Stanford-Binet intelligence scale (N=281), the Wechsler Intelligence Scale for Children – Fourth Edition (WISC-IV)[14] (N=46), the Wechsler Abbreviated Scale of Intelligence – First and Second Editions (WASI-I, WASI-II) (N=338) or the Wechsler Preschool and Primary Scale of Intelligence – Fourth Edition (WPPSI-IV) (N=128).

In ASD cohorts, we did not adjust for age as we adjusted for the type of test used for assessing IQ that already takes into account the age (as well as language level) of the individual. Cognitive scores were derived as follows:

Linear Regression = glm(Cognitive ability ZScore ~ Cognitive Test Type, data= ASD Cohort)
ASD Cohort$residual_Zscore = residuals(Linear Regression)

*Imagen Cohort.* Scores were obtained from the fourth edition of the Wechsler intelligence scale for children (WISC-IV). Deviation NVIQ were available for all participants.

*Generation Scotland Cohort.* The g-factor is based on four cognitive tests measuring processing speed, verbal declarative memory, executive functions and vocabulary. The g-factor represents 42.3% of the observed variance. The g-factor was then transformed to a z-score using the mean of -3.649 x **~**10^-16^ and the SD of 1.3. For more details see Huguet et al., 2021[15]

Sleep.

To date four SSC papers have been published using the Sleep Interview (SSCI), to examine parent reported sleep disturbances in relation to behavioral, medical and cognitive phenotypes, and most recently with common genetic variants[16–19]. Within the SSC cohort, nighttime problems, particularly difficulty going to sleep, are the most frequently reported sleep issue[17]. These nighttime scores have been associated with medical and cognitive problems, like gastrointestinal issues and lower NVIQ. To date, items on the SSCI have been aggregated into subscales and global composite scores, while specific sleep phenotype items have yet to be explored in depth. Unlike previous SSC sleep characterizations, current analyses will be more specific to sleep traits related to insomnia that are reported to be elevated in the ASD population.

**DATA ANALYSIS**

*R 3.6.3 Statistical models packages used:*

| **Regression model** | **Pagckage** | **Function** | **Other** |
| --- | --- | --- | --- |
| Bayesian logistic regression | ‘*arm’* | *bayesglm()* | random effect (1\| FID) **Control for family relations* |
| Ordinal logistic regression | *‘MASS’* | *Polr()* |  |
| Linear regression | ‘stats’ | *lm()* |  |
